# Exploring the possible causal role of the immune response to varicella-zoster virus on multiple traits: a phenome-wide Mendelian randomization study

**DOI:** 10.1101/2022.07.29.22278202

**Authors:** Xinzhu Yu, Artitaya Lophatananon, Krisztina Mekli, Kenneth R. Muir, Hui Guo

## Abstract

**Background:** The immune response to infections could be largely driven by the individual’s genes, especially in the major histocompatibility complex (MHC) region. Varicella-zoster virus (VZV) is a highly communicable pathogen. In addition to infection, the re-activations of VZV can be a potential causal factor for multiple traits. Identification of VZV immune response-related health conditions can therefore help elucidate the aetiology of certain diseases.

**Methods:** A phenome-wide Mendelian randomization study (MR-PheWAS) of anti-VZV Immunoglobulin G (IgG) level with 1,370 traits was conducted to explore the potential causal role of VZV-specific immunity on multiple traits using the UK Biobank cohort. For each trait, we performed Mendelian randomization (MR) analyses using five methods with or without instrumental variables (IVs) in the MHC region.

**Results:** We identified 49 single nucleotide polymorphisms (SNPs) associated with anti-VZV IgG level as IVs, five of which were located in the MHC region. Statistical evidence for a causal effect of anti-VZV IgG level on 75 traits was found in at least three of the MR methods, 60 traits if IVs in the MHC region were removed from MR analysis. With or without the MHC IVs, we found that a higher anti-VZV IgG level led to increased risk of 14 diseases and lower risks of 9 diseases. In addition, anti-VZV IgG level was causally associated with 5 biomarker-related traits.

**Conclusions:** Anti-VZV IgG was causally associated with multiple traits. MHC IVs had a substantial impact on MR causal inference, and therefore, should not be neglected from analysis.

## Background

Varicella-zoster virus (VZV) is a highly communicable pathogen. It is widely present in the general population, with an estimated incidence of 4-4.5 per 1,000 person-years [1]. Primary infections of VZV are commonly seen in children as chickenpox. The virus will then remain latent in the sensory dorsal root ganglion cells and can be reactivated throughout life. The reactivations of the virus may cause damage anywhere on the body, from nerves to the skin [2]. The common manifestations are neurological symptoms, such as radicular pain, itching, and unpleasant sensations. The virus can affect the cranial nerve and cause diseases (e.g., tooth exfoliation [3] and facial palsy [2]). Facial palsy could trigger other symptoms such as tinnitus, hearing loss and nausea. VZV reactivation may also result in retinal necrosis and severe ocular diseases [4]. The virus can directly enter the cerebrospinal fluid causing meningitis [5], or invade the spinal cord and produce myelopathy [6]. It is also possible that the virus travels to arteries to induce vasculopathy like haemorrhagic stroke [7]. Recent studies have shown associations between herpes zoster and other diseases such as depression and anxiety [8]. Therefore, the infection of VZV can have a long-term impact on multiple health conditions.

Humoral immune responses to the same infectious agent vary greatly between individuals. Evidence shows that genetic factors may play a determinant role in the individual antibody responses to a variety of viruses, especially the variants in the major histocompatibility complex (MHC) region [9]. Genes likely affect the humoral immunity response by controlling the serum immunoglobulin levels, seroconversion rates and intensity of antigen-specific immune responses [10]. Therefore, once a host is exposed to the pathogen, their immune response will be affected by genetic factors. Studying the infectious immunity associated genes helps understand the pathologic mechanisms. Immunoglobulin G (IgG) antibody is the most common antibody in the blood and is persistent throughout a person’s life [11]. It can be used as a stable biomarker of lifetime exposure to viruses. Increased level of anti-VZV IgG correlates with VZV reactivation [12]. Previous genome-wide association studies (GWASs) investigated SNPs that influence the antibody responses against VZV infections. While Petars et.al, found no SNPs significantly associated with anti-VZV IgG level in 1,000 healthy individuals [10], five SNPs (rs13197633, rs34073492, rs56401801, rs13204572, rs1048381) were identified in a larger cohort (N= 7,595) from the UK biobank [13].

Mendelian randomization (MR) is a well-established tool for inferring causal relationships. Using SNPs as instrumental variables (IVs), it helps to avoid the issue of unobserved confounding in observational studies. The MHC region is a complex genomic region, in which genes are associated with the risks of many diseases [14]. IVs in this region might violate the MR assumptions due to horizontal pleiotropic effects [15]. However, simply excluding them from MR analysis may lead to biased estimation. In fact, several robust MR approaches have been emerged more recently with the aim to minimize the bias from horizontal pleiotropy and/or outliers. For example, MR-Egger estimation allows a genetic instrument to have a direct effect on the outcome, as long as the direct effect is independent of the instrument strength [16]. The weighted median method provides consistent causal estimates if at least 50% of the weights come from valid IVs [17]. MR-RAPS assumes that the pleiotropic effects are balanced and assigns low weights to outliers when estimating the causal effect [18]. The MR-PRESSO method detects the outliers which will then be removed to obtain robust estimations [19]. Nevertheless, each method has their own strengths and weaknesses and requires specific assumptions. It has been recommended that multiple MR methods can be performed to seek consistent evidence for a causal relationship [20].

This study aimed to explore the causal role that VZV specific immune responses play in the development of multiple traits, by performing phenome-wide MR studies (MR-PheWAS) between VZV titre measurement and 1,370 independent traits using five different MR approaches. This study also explored the impact of the genetic instruments in the MHC region on MR analysis.

## Method

The outline of the study is shown in Figure 1.

**Figure 1.**
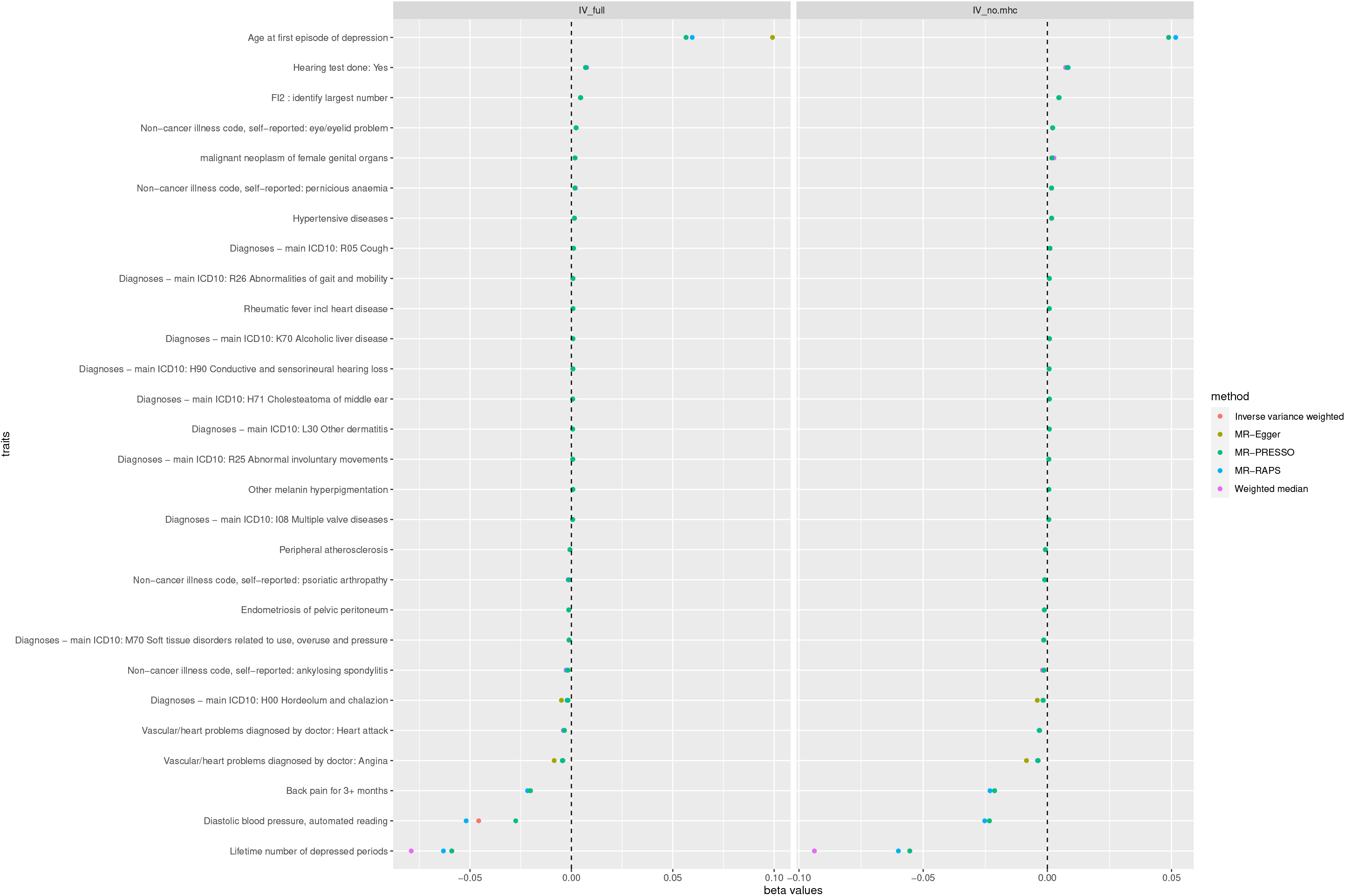
Flowchart of the phenome-wide Mendelian randomization study.

### Identification of Instrumental variables

The UK Biobank is a large-scale prospective database that has recruited more than 500,000 participants aged between 40 and 69. Within the cohort, 10,000 people were randomly selected to undertake the anti-VZV IgG tests [21]. We extracted genotype data and VZV IgG antibody measurement from the UK Biobank locked in March 2021. After excluding individuals with high missing rate (missing rate > 0.02), outlying heterozygosity rate (± 3SD from the mean) and unmatched sex, non-Caucasians, and related individuals, 6,425 individuals were included in the analysis. The anti-VZV IgG titre were measured using fluorescent bead-based multiplex serology technology at serum dilution 1:1,000 [21]. The output data was median fluorescence intensity (MFI) values. The titre data was log 10-transformed due to skewness. We tested association between anti-VZV IgG and each SNP using linear regression model. SNPs with low minor allele frequency (MAF < 0.01), high missing rate (missing rate > 0.02) and low Hardy-Weinberg Equilibrium *P*-value (*P <*1× 10^−6^) were removed for downstream analysis. The SNPs associated with the anti-VZV IgG level were then carried forward for MR analysis. To increase the statistical power, we relaxed the association threshold of *P-*value to 1 × 10^−4^ to include more SNPs in selection of IVs because some of the MR methods used in this study are less prone to weak instrument bias. Clumping was performed on all the neighbouring SNPs (within 10,000kb) to filter out those in linkage disequilibrium (LD) (R^2^ >= 0.01), and only retained the one with the lowest *P*-value. SNPs selected after clumping were mutually independent and used as IVs for MR analysis. Association analysis between SNPs and anti-VZV IgG and SNP clumping were conducted in PLINK v1.9.

### The dataset

We used anti-VZV IgG level as the exposure. The summary statistics (estimated SNP effects and their standard errors) were extracted from our SNP-anti-VZV IgG association analysis. The Neale lab GWASs included 361,194 European descendants from the UK Biobank for over 4,000 traits, from which we only included diseases and biomarker-related traits as the outcomes. The summary statistics of SNP-outcome associations were obtained from the Neale Lab’s GWASs (round 2, http://www.nealelab.is/uk-biobank). We included traits coded from PHESANT, FinnGen and ICD10. Certain diseases such as injuries and chromosomal abnormalities were excluded (details of the criteria code were in Supplementary file 1). We also included the biomarker-related traits, such as symptoms (e.g., knee pain) and body function measurements (e.g., heel density, FEV1). The transient function measurements (e.g., microalbumin in urine, blood cell count, creatinine (quantile)) which fluctuate over time were removed. For continuous traits, we selected rank normalized data. For binary traits, we filtered out those with small number of cases (< 200) to increase the statistical power. For those traits measured in multiple ways, we only report one of them following the criteria of 1) doctor-diagnosed rather than self-reported, 2) the one reflecting general condition rather than period-specific (e.g., general alcohol intake frequency vs alcohol consumption in last few days), 3) for duplicated traits with the same phenotype descriptions, the one with a larger number of cases, less missingness or newer versions were selected. We used single-gender data for gender-related diseases (e.g., female-only GWAS results for ovary-related traits). We used GWAS results of both sexes for all the remaining traits.

### MR analysis

For each trait, we performed two sets of MR tests - including IVs from the MHC region (IV_full_) or not (IV_no.mhc_). The MHC region was defined as chr6:28,477,797-33,448,354 (GRCh37) (The Genome Reference Consortium, https://www.ncbi.nlm.nih.gov/grc). Harmonization was conducted for IVs between exposure and outcome datasets to ensure the same minor allele was used for each IV. MR analyses were conducted using five methods: random effect inverse-variance weighted method [20] and four robust methods including weighted median [17], MR-Egger [16], MR-PRESSO [19], MR-RAPS [18]. We reported a non-zero causal relationship between anti-VZV IgG level and a health condition if there was consistent statistical evidence (*P*-value < 0.05) in at least three out of the five methods and in at least one robust method. MR analyses were implemented using ‘MRPRESSO’, ‘TwoSampleMR’ and ‘MendelianRandomization’ packages in R.

### Results

The mean anti-VZV IgG level (log 10-transformed) and standard deviation are 2.724 and 0.514, ranging from 0 to 4.021 (Supplementary Table S1). We identified 1,421 SNPs associated with anti-VZV IgG, most of which (N = 1,205) were in the MHC region. Details of the results can be found in Supplementary Table S2. Four SNPs (rs13197633, rs13204572, rs1048381 and rs56401801) were also found associated with anti-VZV IgG in previous UK biobank GWAS [13]. Forty-nine independent SNPs (IV_full_) and 44 non-MHC SNPs (IV_no.mhc_) associated with anti-VZV IgG were selected as IVs, respectively, in MR analysis (Table 1). IV_full_ explained 14.5% variation of the anti-VZV IgG level (F-statistic = 935) while IV_no.mhc_ explained 11.4% of the variation (F-statistic = 735). A total of 1,370 traits (846 ordinal, 459 binary and 65 continuous) were included in MR analysis. Supplementary Table S3 lists all these traits.

**Table 1.** Instrumental variables identified by the Genome-wide association scan^1^

| SNP | CHR | Position | Minor allele | Beta | SE | MAF | *P* value | MHC region |
| --- | --- | --- | --- | --- | --- | --- | --- | --- |
| rs749003 | 1 | 118531980 | C | 0.053 | 0.013 | 0.142 | 4.98E-05 | No |
| rs843971 | 1 | 153277423 | T | -0.037 | 0.009 | 0.367 | 7.71E-05 | No |
| rs114925113 | 1 | 170714714 | A | -0.139 | 0.033 | 0.017 | 2.63E-05 | No |
| rs2642449 | 1 | 220985677 | C | -0.060 | 0.013 | 0.139 | 3.06E-06 | No |
| rs708108 | 1 | 228189855 | T | -0.037 | 0.009 | 0.402 | 8.41E-05 | No |
| rs74482711 | 2 | 127357322 | G | -0.067 | 0.016 | 0.092 | 3.74E-05 | No |
| rs34499171 | 2 | 129591736 | T | 0.045 | 0.011 | 0.192 | 7.62E-05 | No |
| rs61731470 | 2 | 211085491 | A | 0.112 | 0.028 | 0.028 | 5.96E-05 | No |
| rs12471761 | 2 | 241310093 | G | -0.110 | 0.028 | 0.028 | 7.47E-05 | No |
| rs17495909 | 3 | 1323130 | C | -0.097 | 0.022 | 0.042 | 1.22E-05 | No |
| rs809764 | 3 | 61958023 | G | -0.039 | 0.010 | 0.327 | 5.27E-05 | No |
| rs116664692 | 3 | 62271345 | G | 0.180 | 0.046 | 0.010 | 8.90E-05 | No |
| rs11706859 | 3 | 100713091 | C | -0.137 | 0.034 | 0.019 | 7.03E-05 | No |
| rs35573656 | 3 | 126612552 | C | -0.073 | 0.018 | 0.072 | 3.00E-05 | No |
| rs73112967 | 4 | 27778474 | C | -0.083 | 0.020 | 0.055 | 5.44E-05 | No |
| rs156572 | 4 | 155739138 | T | -0.109 | 0.025 | 0.036 | 1.23E-05 | No |
| rs72696744 | 4 | 163745634 | T | 0.117 | 0.029 | 0.025 | 4.75E-05 | No |
| rs1395479 | 4 | 178318191 | A | -0.043 | 0.010 | 0.264 | 2.68E-05 | No |

<sup>1</sup> MAF: Minor allele frequency

|  |  |  |  |  |  |  |  |  |
| --- | --- | --- | --- | --- | --- | --- | --- | --- |
| rs78827810 | 4 | 186683778 | G | -0.115 | 0.029 | 0.026 | 8.06E-05 | No |
| rs56244403 | 6 | 24017867 | T | -0.038 | 0.010 | 0.383 | 8.13E-05 | No |
| rs12193110 | 6 | 29937104 | T | -0.065 | 0.010 | 0.299 | 8.83E-11 | Yes |
| rs17200698 | 6 | 31483700 | T | -0.087 | 0.022 | 0.049 | 5.14E-05 | Yes |
| rs116462901 | 6 | 31488532 | C | -0.138 | 0.032 | 0.020 | 2.17E-05 | Yes |
| rs3129888 | 6 | 32411726 | G | -0.058 | 0.011 | 0.194 | 2.66E-07 | Yes |
| rs1048381 | 6 | 32610445 | A | 0.116 | 0.012 | 0.171 | 9.28E-23 | Yes |
| rs80018618 | 6 | 37337516 | A | 0.161 | 0.039 | 0.013 | 4.04E-05 | No |
| rs41265357 | 6 | 71238178 | G | -0.186 | 0.047 | 0.010 | 7.06E-05 | No |
| rs2180346 | 6 | 168944598 | C | 0.072 | 0.015 | 0.106 | 9.25E-07 | No |
| rs72695580 | 9 | 6632950 | G | 0.170 | 0.042 | 0.014 | 6.02E-05 | No |
| rs11139344 | 9 | 84239037 | C | 0.042 | 0.010 | 0.268 | 3.49E-05 | No |
| rs116919233 | 9 | 119991655 | T | -0.123 | 0.031 | 0.021 | 8.77E-05 | No |
| rs1072523 | 10 | 28819406 | A | 0.036 | 0.009 | 0.492 | 8.63E-05 | No |
| rs3759012 | 11 | 118901262 | T | 0.036 | 0.009 | 0.386 | 8.99E-05 | No |
| rs11609865 | 12 | 105952417 | C | -0.073 | 0.018 | 0.068 | 5.83E-05 | No |
| rs116950913 | 12 | 132889076 | A | -0.101 | 0.025 | 0.038 | 4.66E-05 | No |
| rs75782087 | 13 | 38822878 | T | -0.096 | 0.023 | 0.037 | 3.94E-05 | No |
| rs111259346 | 13 | 39377730 | G | -0.092 | 0.024 | 0.034 | 9.86E-05 | No |
| rs73241676 | 13 | 82454247 | G | 0.055 | 0.014 | 0.131 | 5.36E-05 | No |
| rs72740509 | 15 | 56533519 | T | -0.055 | 0.014 | 0.123 | 6.59E-05 | No |
| rs9747501 | 17 | 2739061 | A | 0.043 | 0.010 | 0.260 | 3.62E-05 | No |
| rs12948236 | 17 | 65633567 | C | 0.037 | 0.009 | 0.426 | 6.97E-05 | No |
| rs6507201 | 18 | 35016217 | A | 0.038 | 0.009 | 0.359 | 6.26E-05 | No |
| rs319445 | 18 | 55360524 | A | 0.055 | 0.014 | 0.122 | 8.86E-05 | No |
| rs622989 | 19 | 7574308 | C | 0.127 | 0.032 | 0.020 | 6.79E-05 | No |
| rs34335847 | 19 | 14705499 | A | -0.061 | 0.015 | 0.102 | 4.25E-05 | No |
| rs45465702 | 20 | 33320571 | C | -0.050 | 0.012 | 0.184 | 2.04E-05 | No |
| rs78492969 | 20 | 49248213 | T | -0.101 | 0.024 | 0.037 | 2.39E-05 | No |
| rs62222450 | 21 | 40471448 | T | 0.073 | 0.018 | 0.063 | 6.07E-05 | No |
| rs117884976 | 22 | 28107887 | T | -0.112 | 0.029 | 0.024 | 9.00E-05 | No |

### MR results

Seventy-five and 60 traits were causally associated with anti-VZV IgG level, respectively, with and without IVs from the MHC region (Supplementary Table S4). For each trait, although the estimated causal effects varied, the direction of the association was the same across different MR methods. Whether including MHC IVs or not in MR, causal relationships were consistently found between anti-VZV IgG and 28 traits (Figure 2), and the directions of the causal effects were concordant (Table 2).

**Figure 2.**
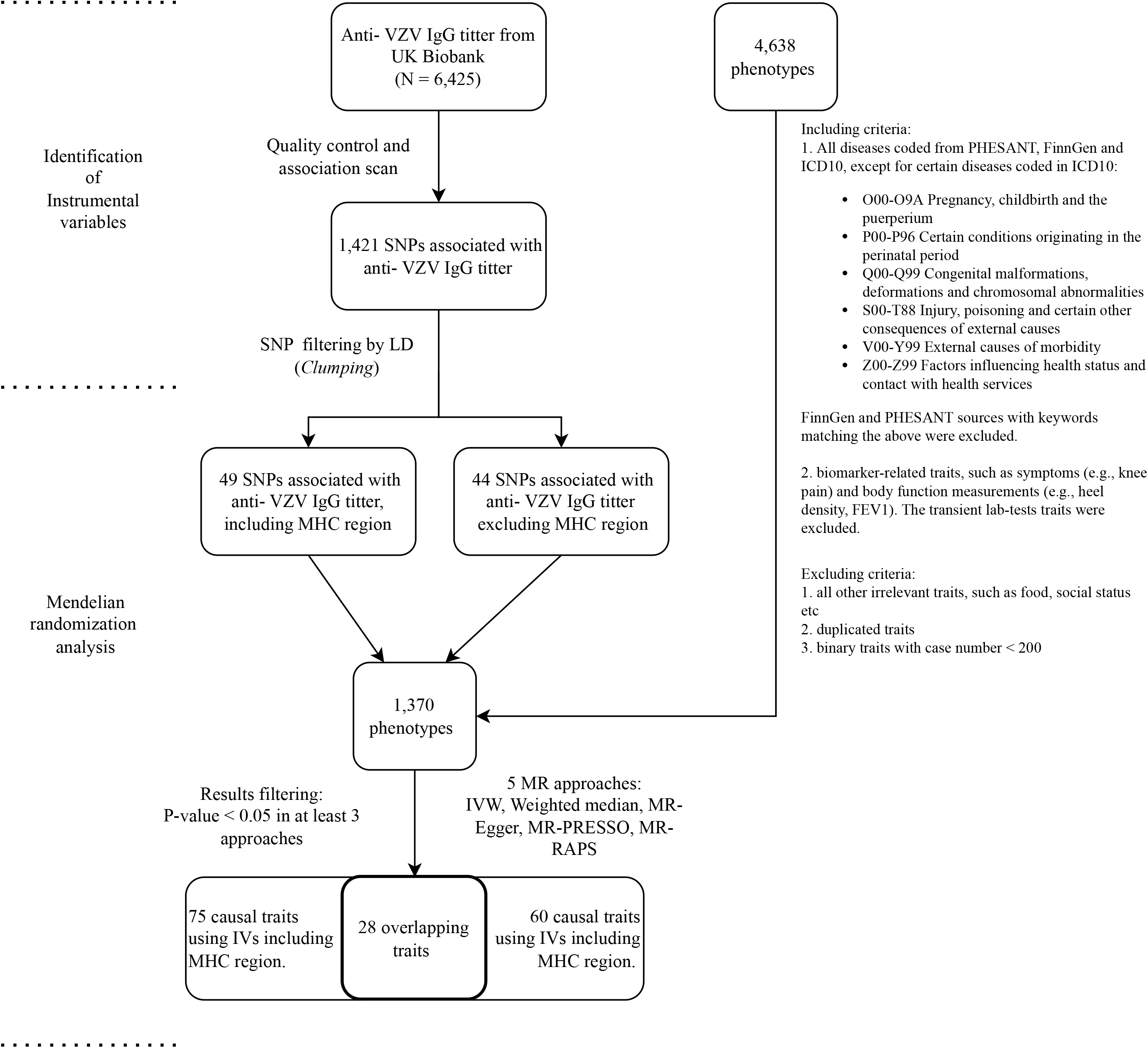
Scatterplot of the causal coefficients (beta values) between anti-VZV IgG level and traits that have significant result (*P* <0.05) in at least three Mendelian randomization (MR) approaches, in both IV strategies (IV_full_ and IV_no.mhc_). The figure on the left is the MR results with IVs including MHC region (IV_full_). The figure on the right is the MR results with IVs excluding MHC region (IV_no.mhc_). Y-axis represents different traits. X-axis is the scale of beta-value. Each dot color represents different MR methods (e.g., orange for inverse variance weighted method).

**Table 2.** Phenome-wide MR results of traits that have significant results (*P* < 0.05) in at least three MR approaches in both IV strategies (IV_full_ and IVno._MHC_)^2^

| Phenotype description | Sample size | Controls | Cases | Source | IV | Beta |  |  |  |  |
| --- | --- | --- | --- | --- | --- | --- | --- | --- | --- | --- |
|  |  |  |  |  |  | Weighted median | IVW | MR-RAPS | MR-PRESSO | MR-Egger |
| Age at first episode of depression | 61,033 | - | - | PHESANT | IV<sub>full</sub> | - | 0.057 | 0.060 | 0.057 | 0.099 |
|  |  |  |  |  | IV<sub>no.mhc</sub> | - | 0.049 | 0.052 | 0.049 | - |
| Back pain for 3+ months | 90,555 | 28,500 | 62,055 | PHESANT | IV<sub>full</sub> | - | -0.020 | -0.022 | -0.020 | - |
|  |  |  |  |  | IV<sub>no.mhc</sub> | - | -0.021 | -0.023 | -0.021 | - |
| Diagnoses - main ICD10: H00 Hordeolum and chalazion | 361,194 | 359,727 | 1,467 | ICD10 | IV<sub>full</sub> | -0.002 | -0.002 | -0.002 | -0.002 | -0.005 |
|  |  |  |  |  | IV<sub>no.mhc</sub> | - | -0.002 | -0.002 | -0.002 | -0.004 |
| Diagnoses - main ICD10: H71 Cholesteatoma of middle ear | 361,194 | 360,864 | 330 | ICD10 | IV<sub>full</sub> | - | 0.001 | 0.001 | 0.001 | - |
|  |  |  |  |  | IV<sub>no.mhc</sub> | - | 0.001 | 0.001 | 0.001 | - |
| Diagnoses - main ICD10: H90 Conductive and sensorineural hearing loss | 361,194 | 360,912 | 282 | ICD10 | IV<sub>full</sub> | - | 0.001 | 0.001 | 0.001 | - |
|  |  |  |  |  | IV<sub>no.mhc</sub> | - | 0.001 | 0.001 | 0.001 | - |
| Diagnoses - main ICD10: I08 Multiple valve diseases | 361,194 | 360,944 | 250 | ICD10 | IV<sub>full</sub> | - | 0.001 | 0.001 | 0.001 | - |
|  |  |  |  |  | IV<sub>no.mhc</sub> | - | 0.001 | 0.001 | 0.001 | - |
| Diagnoses - main ICD10: K70 Alcoholic | 361,194 | 360,850 | 344 | ICD10 | IV<sub>full</sub> | - | 0.001 | 0.001 | 0.001 | - |

<sup>2</sup> IV<sub>full</sub>: instrumental variables including SNPs in the MHC region IV<sub>MHC</sub>: instrumental variables excluding SNPs in the MHC region -: not applicable or results not significant IVW: inverse-variance weighted method

|  |  |  |  |  |  |  |  |  |  |  |
| --- | --- | --- | --- | --- | --- | --- | --- | --- | --- | --- |
| liver disease |  |  |  |  | IV<sub>no.mhc</sub> | 0.001 | 0.001 | 0.001 | 0.001 | - |
| Diagnoses - main ICD10: L30 Other dermatitis | 361,194 | 360,860 | 334 | ICD10 | IV<sub>full</sub> | - | 0.001 | 0.001 | 0.001 | - |
|  |  |  |  |  | IV<sub>no.mhc</sub> | - | 0.001 | 0.001 | 0.001 | - |
| Diagnoses - main ICD10: M70 Soft tissue disorders related to use overuse and pressure | 361,194 | 360,115 | 1,079 | ICD10 | IV<sub>full</sub> | - | -0.001 | -0.001 | -0.001 | - |
|  |  |  |  |  | IV<sub>no.mhc</sub> | -0.002 | -0.001 | -0.002 | -0.001 | - |
| Diagnoses - main ICD10: R05 Cough | 361,194 | 360,210 | 984 | ICD10 | IV<sub>full</sub> | - | 0.001 | 0.001 | 0.001 | - |
|  |  |  |  |  | IV<sub>no.mhc</sub> | - | 0.001 | 0.001 | 0.001 | - |
| Diagnoses - main ICD10: R25 Abnormal involuntary movements | 361,194 | 360,961 | 233 | ICD10 | IV<sub>full</sub> | 0.001 | 0.001 | 0.001 | 0.001 | - |
|  |  |  |  |  | IV<sub>no.mhc</sub> | - | 0.001 | 0.001 | 0.001 | - |
| Diagnoses - main ICD10: R26 Abnormalities of gait and mobility | 361,194 | 360,864 | 330 | ICD10 | IV<sub>full</sub> | - | 0.001 | 0.001 | 0.001 | - |
|  |  |  |  |  | IV<sub>no.mhc</sub> | - | 0.001 | 0.001 | 0.001 | - |
| Diastolic blood pressure automated reading | 340,162 | - | - | PHESANT | IV<sub>full</sub> | - | -0.046 | -0.052 | -0.027 | - |
|  |  |  |  |  | IV<sub>no.mhc</sub> | - | -0.023 | -0.025 | -0.023 | - |
| Endometriosis of pelvic peritoneum | 194,174 | 193,886 | 288 | FinnGen | IV<sub>full</sub> | - | -0.001 | -0.001 | -0.001 | - |
|  |  |  |  |  | IV<sub>no.mhc</sub> | - | -0.001 | -0.001 | -0.001 | - |
| FI2 : identify largest number | 116,869 | 950 | 115,919 | PHESANT | IV<sub>full</sub> | - | 0.004 | 0.005 | 0.004 | - |
|  |  |  |  |  | IV<sub>no.mhc</sub> | - | 0.005 | 0.005 | 0.005 | - |
| Hearing test done: Yes | 119,880 | 2,675 | 117,205 | PHESANT | IV<sub>full</sub> | 0.008 | 0.007 | 0.007 | 0.007 | - |
|  |  |  |  |  | IV<sub>no.mhc</sub> | 0.007 | 0.008 | 0.008 | 0.008 | - |
| Hypertensive diseases | 361,194 | 359,881 | 1,313 | FinnGen | IV<sub>full</sub> | - | 0.001 | 0.002 | 0.001 | - |
|  |  |  |  |  | IV<sub>no.mhc</sub> | - | 0.002 | 0.002 | 0.002 | - |
| Lifetime number of depressed periods | 53,691 | - | - | PHESANT | IV<sub>full</sub> | -0.079 | -0.059 | -0.063 | -0.059 | - |
|  |  |  |  |  | IV<sub>no.mhc</sub> | -0.094 | -0.055 | -0.060 | -0.055 | - |
| Malignant neoplasm of female genital organs | 361,194 | 358,591 | 2,603 | FinnGen | IV<sub>full</sub> | - | 0.002 | 0.002 | 0.002 | - |
|  |  |  |  |  | IV<sub>no.mhc</sub> | 0.003 | 0.002 | 0.002 | 0.002 | - |
| Non-cancer illness code self-reported: ankylosing spondylitis | 361,141 | 360,103 | 1,038 | PHESANT | IV<sub>full</sub> | -0.003 | -0.002 | -0.002 | -0.002 | - |
|  |  |  |  |  | IV<sub>no.mhc</sub> | -0.002 | -0.001 | -0.001 | -0.001 | - |
| Non-cancer illness code self-reported: eye/eyelid problem | 361,141 | 358,301 | 2,840 | PHESANT | IV<sub>full</sub> | - | 0.002 | 0.002 | 0.002 | - |
|  |  |  |  |  | IV<sub>no.mhc</sub> | - | 0.002 | 0.002 | 0.002 | - |
| Non-cancer illness code self-reported: pernicious anaemia | 361,141 | 360,032 | 1,109 | PHESANT | IV<sub>full</sub> | 0.002 | 0.002 | 0.002 | 0.002 | - |
|  |  |  |  |  | IV<sub>no.mhc</sub> | - | 0.002 | 0.002 | 0.002 | - |
| Non-cancer illness code self-reported: psoriatic arthropathy | 361,141 | 360,429 | 712 | PHESANT | IV<sub>full</sub> | -0.002 | -0.001 | -0.001 | -0.001 | - |
|  |  |  |  |  | IV<sub>no.mhc</sub> | - | -0.001 | -0.001 | -0.001 | - |
| Other melanin hyperpigmentation | 361,194 | 360,876 | 318 | FinnGen | IV<sub>full</sub> | - | 0.001 | 0.001 | 0.001 | - |
|  |  |  |  |  | IV<sub>no.mhc</sub> | - | 0.001 | 0.001 | 0.001 | - |
| Peripheral atherosclerosis | 361,194 | 360,690 | 504 | FinnGen | IV<sub>full</sub> | - | -0.001 | -0.001 | -0.001 | - |
|  |  |  |  |  | IV<sub>no.mhc</sub> | - | -0.001 | -0.001 | -0.001 | - |
| Rheumatic fever incl heart disease | 361,194 | 360,760 | 434 | FinnGen | IV<sub>full</sub> | - | 0.001 | 0.001 | 0.001 | - |
|  |  |  |  |  | IV<sub>no.mhc</sub> | - | 0.001 | 0.001 | 0.001 | - |
| Vascular/heart problems diagnosed by doctor: Angina | 360,420 | 349,048 | 11,372 | PHESANT | IV<sub>full</sub> | - | -0.004 | -0.005 | -0.004 | -0.008 |
|  |  |  |  |  | IV<sub>no.mhc</sub> | - | -0.004 | -0.004 | -0.004 | -0.008 |
| Vascular/heart problems diagnosed by doctor: Heart attack | 360,420 | 352,132 | 8,288 | PHESANT | IV<sub>full</sub> | -0.004 | -0.003 | -0.004 | -0.003 | - |
|  |  |  |  |  | IV<sub>no.mhc</sub> | - | -0.003 | -0.003 | -0.003 | - |

Among the 28 traits, 14 were positively causally associated with anti-VZV IgG level, including eye/eyelid problem, pernicious anaemia, malignant neoplasm of female genital organs, hypertensive diseases, cough, conductive and sensorineural hearing loss, rheumatic fever, abnormalities of gait and mobility, alcoholic liver disease, other melanin hyperpigmentation, abnormal involuntary movements, multiple valve diseases, cholesteatoma of middle ear, and other dermatitis. Elevated anti-VZV IgG level in the body increased the risks of these diseases. Anti-VZV IgG level was also positively causally associated with age at first episode of depression, completing hearing test and identifying number in the fluid intelligence test. The higher the anti-VZV IgG level in the body, the older estimated age at depression and the higher likelihood of completing hearing tests and better performance in one of the intelligent tests would be.

We found negative causal associations between anti-VZV IgG level and 9 diseases, including peripheral atherosclerosis, soft tissue disorders related to overuse and pressure, psoriatic arthropathy, endometriosis of pelvic peritoneum, ankylosing spondylitis, hordeolum and chalazion, heart attack, angina, and back pain. It was shown that high anti-VZV IgG level reduced the risk of these diseases. Anti-VZV IgG level was also negatively causally associated with lifetime number of depressed periods and diastolic blood pressure (DBP). The results showed high anti-VZV IgG level associated with a smaller number of expressed periods and lower DBP.

In addition, we found 79 traits showed inconsistent causal associations with anti-VZV IgG using IV_full_ and IV_no.mhc_ separately in MR analysis. Forty-seven traits were significantly associated with anti-VZV IgG level using IV_full_ only (Supplementary Figure S1A), whereas 32 traits showed evidence of causal relationship with anti-VZV IgG using IV_no.mhc_ (Supplementary Figure S1B), but not IV_full_.

Hordeolum and chalazion was causally associated with anti-VZV IgG level in all five MR methods using IV_full_. However, when IV_no.mhc_ were used in MR, weighted median method merely showed a marginally significant result (*P* = 0.052). The causal evidence between hordeolum and chalazion and anti-VZV IgG level was consistent across the MR methods. Ankylosing spondylitis, completing hearing test and lifetime number of depressed periods were significantly associated with anti-VZV IgG level in four MR approaches except for MR-Egger in both sets of IVs. Angina was significantly associated with anti-VZV IgG level in four MR approaches except for weighted median method in both sets of IVs. The evidence of causal relationship between anti-VZV IgG level and other traits were less consistent across the MR methods or IVs sets, but they all showed evidence for a causal relationship in at least three MR methods.

## Discussion

Our MR results suggested anti-VZV IgG level is positively causally associated with eye/ eyelid problems. VZV reactivation can affect nerve that innervate ocular and periocular structures, named herpes zoster ophthalmicus (HZO). HZO can lead to a variety of ocular diseases such as keratitis, retinitis, and glaucoma etc [22]. As VZV could spread to the central nervous system (CNS) and cause CNS diseases accordingly [23], it is not surprising to see a positive causal relationship between it and abnormal involuntary movements. Although rare, cases have been reported for VZV causing involuntary movement [24]. Higher anti-VZV IgG level was also causally associated with a smaller number of time with depression and experiencing depression at older age. The results suggested anti-VZV IgG level to be a protective causal factor for depression. It was in concordance with previous finding of negative associations between depression and the VZV-specific cell-mediated immunity (VZV-CMI) levels [25]. We also found the anti-VZV IgG was potentially a causal risk factor of conductive and sensorineural hearing loss and cholesteatoma of the middle ear. Cholesteatoma is commonly caused by repeated infections or inflammation in the middle ear, the consequences likely to be caused by VZV reactivation. The disorder may further lead to hearing loss [26]. Moreover, reactivations of VZV can cause severe damage to the facial nerve, leading to conditions like Ramsay Hunt syndrome, and may subsequently, result in loss of facial functions, including hearing loss [27]. There were cases of sudden sensorineural hearing loss directly caused by VZV [28]. These findings all suggest that exposure to VZV is a potential causal risk factor for ear diseases. Although we also found higher anti-VZV IgG level associated with higher completeness of hearing tests, which seemed inconsistent with the other evidence. The competence of hearing tests was less persuasive comparing to other traits, as it could be affected by many factors such as environmental or mental issues.

This could also be true for one of the cognitive tests (FI2) and DBP. A negative causal relationship between these traits and anti-VZV IgG level was observed in our study. It is known VZV could spread to the CNS and cause CNS diseases, which will possibly lead to brain damages or cognitive impairment [29]. As a positive causal relationship between anti-VZV IgG level and hypertension was detected, the result of transient BP reading was less persuasive as it could be affected by many factors such as the antihypertensive medicine.

Discussion about the relationship between VZV reactivations and cardiovascular diseases have been long-standing. The virus could travel through ganglia to arteries and cause VZV vasculopathy. It has been found that VZV could cause damage to the intracranial circulation, and lead to diseases like stroke [30]. There were suspect of VZV also being risk factors for cardiovascular diseases [31]. But no evidence that it could enter aortas or temporal arteries have been found [32]. Here we detected a negative causal relationship between anti-VZV IgG level and peripheral atherosclerosis, heart attack and angina. We also found anti-VZV IgG level to be positively causally associated with hypertension. Both hypertension and atherosclerosis are well known risk factors for cardiovascular diseases. The different directions of the causal results may provide us with the different causal mechanisms from VZV infections to cardiovascular events. One of the possible pathways from VZV infections to cardiovascular diseases were through rheumatic fever and valve disease. We found that high anti-VZV IgG level increased the risk for rheumatic fever and multiple valve diseases. Multiple valve diseases can be driven by infections. Rheumatic fever is an auto-immune condition that could also damage the heart valves and lead to rheumatic heart disease [33].

The correlation between the incidence of herpes zoster reactivations and autoimmune diseases have also been reported previously [34, 35]. Other than rheumatic fever, we also showed that VZV immune response was causally associated with several auto-immune and auto-inflammation conditions including pernicious anaemia, psoriatic arthropathy, ankylosing spondylitis, and the soft tissue disorders related to overuse and pressure. Pernicious anaemia is an autoimmune disease that affects the stomach against absorbing vitamin B12. Psoriatic arthropathy refers to diseases caused by the autoimmune responses attacking the joint. The arthritis could then lead to abnormalities of gait and mobility. Ankylosing spondylitis is a chronic inflammatory condition that affects spine. The soft tissue disorders related to overuse and pressure in ICD 10 code included multiple auto-inflammatory diseases such as bursitis and synovitis. Except for pernicious anaemia and abnormalities of gait and mobility, anti-VZV IgG level were negatively associated with psoriatic arthropathy, ankylosing spondylitis, and the soft tissue disorders related to overuse and pressure. Thus, the relationship among these traits appeared to be complicated. The virus itself might increase the risk of flares in some auto-immune diseases like systemic lupus erythematosus (SLE) [36]. In another way, herpes zosters are more likely to reactivate in these patients as they might take immunosuppressive medications [37]. However, the immunosuppressive treatment also decreases the antibody levels in one’s body [38]. Therefore, interactions should be considered in future studies.

In our study, we found evidence that high anti-VZV IgG level causally increase the risk of malignant neoplasm of female genital organs. Herpes zoster had been linked to an increased risk of cancer [39], but no evidence for specific cancer type were reported. We also found anti-VZV IgG level causally associated with endometriosis of pelvic peritoneum and alcohol liver disease. The two findings also lack related evidence in the literature. Cough, back pain, other dermatitis, and other hyperpigmentation were also identified to have causal relationship with anti-VZV IgG level. However, the definitions of these traits were rather vague. More detailed investigations into the definitions were important before linking valid causal relationships to them.

Interestingly, some traits that have known evidence of causal association with anti-VZV IgG level such as stroke [30] and SLE [36] were detected in MR analysis with full (IV_full_) or partial sets of instruments (IV_no.mhc_). We detected evidence of anti-VZV IgG level to be causally associated with cancer such as sarcoidosis, neoplasm of the ovary and neoplasms of skin (Supplementary Table S4) with IV_no.mhc_ only. The same relationships were observed for arthritis-related traits, such as hallux rigidus and carpal tunnel syndrome. However, the causal relationship was questionable as they were not detectable if we include IVs on the MHC region.

The MHC region is a complex genomic region due to its linkage disequilibrium and its roles in multiple diseases [40]. Some researchers ignored this region in their MR studies [41], but concerns have been raised about possible pleiotropy when using MHC instruments. Several MR studies removed MHC SNPs from the IV list to minimize the risk of assumption violations [15, 42]. However, MHC SNPs often have strong associations with the exposure, especially for infection-related traits [13]. Simply removing them might introduce weak instrument bias. Here, we explored the impact of MHC instruments on MR analysis. Surprisingly, we found evidence of inconsistency in the MR results introduced by MHC IVs. The heterogeneity of the results between two IV strategies confirmed doubts on using MHC IVs. Simply ignoring the effects of MHC variants could introduce distorted associations into the MR analysis. However, naively removing them from IV list risks losing valid IVs and masking the true causal relationship. A more robust way might be looking at the overlapped results between different IV strategies. Or performing sensitivity analysis that removes MHC IVs for robust estimations.

Previous GWAS identified five SNPs associated with anti-VZV IgG level [13]. Our genome-wide association results was consistent with the previous GWAS in terms of rs13197633, rs13204572, rs1048381 and rs56401801. They have the same direction of effect, and all passed the *P*-value thresholds. Rs34073492, the other SNP identified in previous GWAS, failed at our QC (MAF > 0.02). In addition, we identified more potentially relevant SNPs by relaxing *P*-value threshold of association, which added information on the genetic contribution to the humoral immunity towards VZV.

The anti-VZV IgG level in our study population were not likely driven by vaccination. This is because the age group enrolled in the UK national program for shingles vaccination is between 70-79 years [43], while the enrolled ages for the UK biobank cohort are 40-69. Apart from vaccination, the anti-VZV IgG level could still be driven by many factors, which make the interpretation of the measurement difficult. We used anti-VZV IgG level to represent both seropositivity of VZV and the lifetime exposure to viruses. High anti-VZV IgG level was expected to represent high level of VZV reactivations or immune responses to VZV. However, except for blocking infections, antibodies may have other meaningful functions to the host. It might work in collaboration with cells of the immune system for a more effective protest against pathogens or contribute to the immunity with different unknown mechanisms [44, 45]. Therefore, a high anti-VZV IgG level may also indicate a healthy immune function. This might explain the negative causal association we identified between anti-VZV IgG level and hordeolum and chalazion. The two ocular conditions often caused by infections. A host with healthier immune functions might produce higher amounts of antibodies and therefore be resist of other infections.

## Conclusions

Our phenome-wide MR analysis found anti-VZV IgG level to be a causal risk factor of 28 traits. The findings helped gain new insights into the causal roles of VZV specific immunity in many diseases. While some evidence could be explained in biological ways, others require further investigations. When selecting IVs for MR analysis, researchers should treat MHC SNPs with caution. We recommend that researchers consider using instruments with and without those from the MHC regions, which will help better understand the specific role the MHC region in their MR studies.

## Data Availability

Publicly available data from the UK Biobank study was analysed in this study. The datasets are available to researchers through an open application via https://www.ukbiobank.ac.uk/register-apply/. Publicly available GWAS summary statistics used in this study is available in http://www.nealelab.is/uk-biobank.

## List of abbreviations

BP: blood pressure
CNS: central nervous system
GWAS: genome-wide association study
HZO: herpes zoster ophthalmicus
IgG: immunoglobulin G
LD: linkage disequilibrium
MAF: minor allele frequency
MHC: histocompatibility complex
MR-PheWAS: mendelian randomization phenome-wide association study
SLE: systemic lupus erythematosus
SNP: single nucleotide polymorphisms
VZV: varicella-zoster virus

## Declarations

### Consent for publication

The work described has not been published before. The manuscript is approved by all co-authors.

### Authors’ contributions

XY performed the phenome-wide Mendelian randomization analysis and was a major contributor in writing the manuscript. KM performed the genome-wide association scan. All authors read and approved the final manuscript.

## Acknowledgements

We would like to thank the UK Biobank participants and staff. This study used the UK Biobank application number 5864. We also thank the NHGRI-EBI and Neale lab for making GWAS results publicly accessible.

## Supplementary files

Supplementary Table S1. Descriptive table of anti-VZV IgG level

Table S2. SNPs associated with anti-VZV IgG level identified by the Genome-wide association scan

Table S3. Trait list included in the phenome-wide Mendelian randomization analysis

Table S4. All significant results from the phenome-wide Mendelian randomization analyses (*P* value <0.05) in at least three MR approaches.

Table S5. Full results of the phenome-wide Mendelian randomization analyses of 1,370 traits

Supplementary Documentation 1. Traits selection inclusion and exclusion criteria for the phenome-wide Mendelian randomization study

Figure S1. Scatterplot of the causal coefficients (beta values) between anti-VZV IgG level and traits that have significant result (*P* <0.05) in at least three Mendelian randomization (MR) approaches, only with IV_full_ or IV_no.mhc_. The figure on the left is the MR results with IVs including MHC region (IV_full_). The figure on the right is the MR results with IVs excluding MHC region (IV_no.mhc_). Y-axis represents different traits. X-axis is the scale of beta-value. Each dot color represents different MR methods (e.g., orange for inverse variance weighted method).

