## Supplementary figures and images for "Exploring the possible causal role of the immune response to varicella-zoster virus on multiple traits: a phenome-wide Mendelian randomization study"

### Supplementary Figure

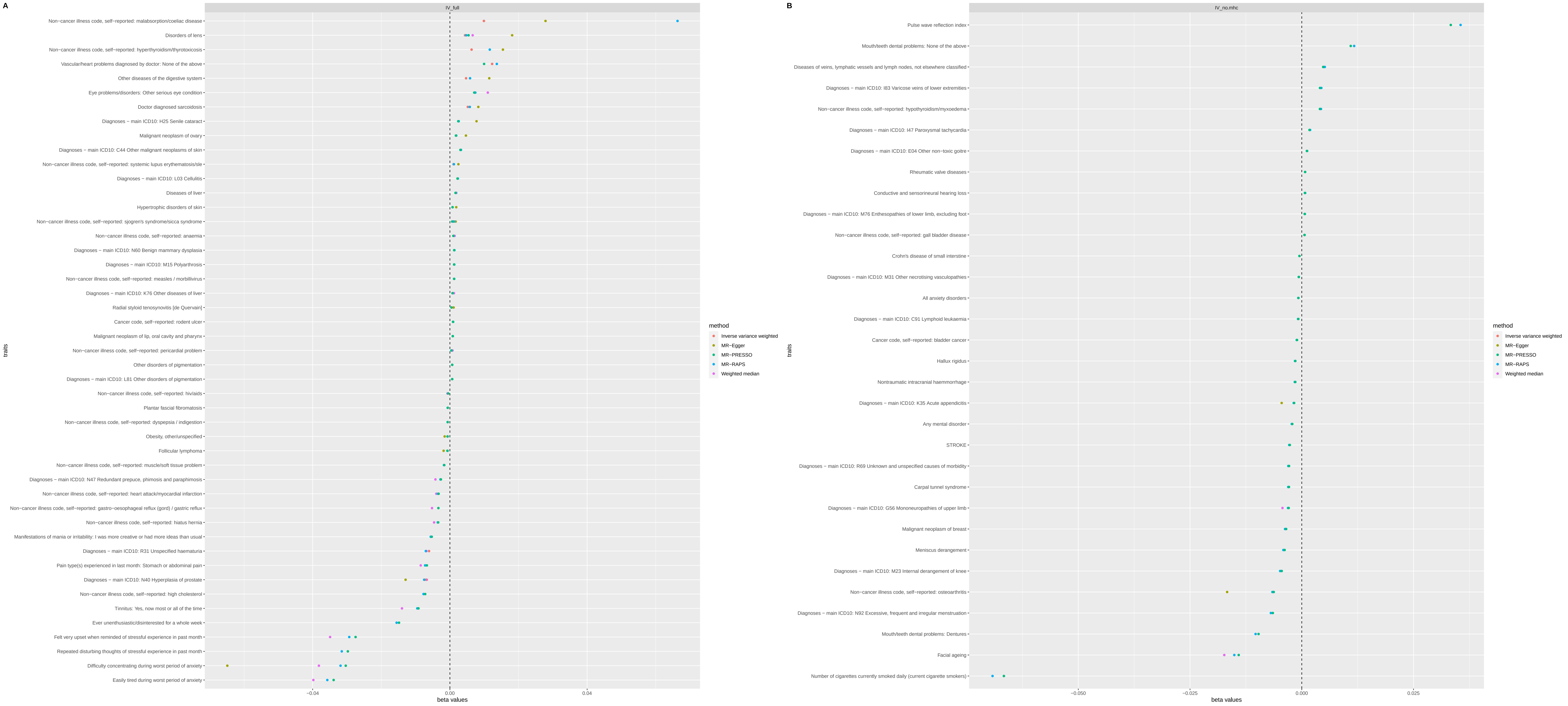
