## Supplementary file 1 for "Exploring the possible causal role of the immune response to varicella-zoster virus on multiple traits: a phenome-wide Mendelian randomization study"

**Traits including and excluding criteria for the phenome-wide Mendelian randomization study**

1. We include disease codes from PHESANT, FinnGen and ICD10 sources. Certain diseases such as injuries, chromosomal abnormalities and gender-determined diseases are excluded. For example, the following codes are excluded from ICD-10 sources:

O00-O9A	Pregnancy, childbirth and the puerperium

P00-P96	Certain conditions originating in the perinatal period

Q00-Q99	Congenital malformations, deformations and chromosomal abnormalities

S00-T88	Injury, poisoning and certain other consequences of external causes

V00-Y99	External causes of morbidity

Z00-Z99	Factors influencing health status and contact with health services

The similar traits coded from FinnGen and PHESANT were also removed.

2. biomarkers, symptoms, signs

We also included the biomarker-related traits, such as symptoms (e.g., knee pain) and body function measurements (e.g., heel density, FEV1). The transient function measurements (e.g., lab-test, Microalbumin in urine, blood cell count, creatinine (quantile)) which cannot represent the long-term health were removed.

2.1 signs (e.g., snoring, knee pain)

2.2 symptoms (e.g., weight change during anxiety/ depression)

2.3 biomarkers

2.4 function measurement (e.g., FEV1)

2.5 disease diagnosed age (e.g., age hypertension diagnosed at)

2.6 exposure (e.g., smoked/drink, only alcohol/tobacco dependency were included)

2.7 physical measurement (e.g., heel bone mineral density, BMI)

3. All other irrelevant traits were removed from the downstream analysis. The full trait list that was included in the study can be found in the supplementary tableS3.

3.1 food (e.g., milk consumption, coffee intake)

3.2 activity (e.g., exercise)

3.3 environment/social status（e.g., home location/pollution, employment）

3.4 experience (e.g., year lived in a region)

3.5 irrelevant signs, measurement (e.g., hair colour, skin colour, arm length)

3.6 treatment (e.g., medication, surgeries)

3.7 time to complete (time to complete the test)

3.8 illness of others (e.g., Alzheimer's of mother)

3.9 reasons (e.g., reasons for doing things)
